# The Overlooked Role of Specimen Preparation in Bolstering Deep Learning-Enhanced Spatial Transcriptomics Workflows

**DOI:** 10.1101/2023.10.09.23296700

**Authors:** Michael Y. Fatemi, Yunrui Lu, Alos B. Diallo, Gokul Srinivasan, Zarif L. Azher, Brock C. Christensen, Lucas A. Salas, Gregory J. Tsongalis, Scott M. Palisoul, Laurent Perreard, Fred W. Kolling, Louis J. Vaickus, Joshua J. Levy

## Abstract

The application of deep learning methods to spatial transcriptomics has shown promise in unraveling the complex relationships between gene expression patterns and tissue architecture as they pertain to various pathological conditions. Deep learning methods that can infer gene expression patterns directly from tissue histomorphology can expand the capability to discern spatial molecular markers within tissue slides. However, current methods utilizing these techniques are plagued by substantial variability in tissue preparation and characteristics, which can hinder the broader adoption of these tools. Furthermore, training deep learning models using spatial transcriptomics on small study cohorts remains a costly endeavor. Necessitating novel tissue preparation processes enhance assay reliability, resolution, and scalability. This study investigated the impact of an enhanced specimen processing workflow for facilitating a deep learning-based spatial transcriptomics assessment. The enhanced workflow leveraged the flexibility of the Visium CytAssist assay to permit automated H&E staining (e.g., Leica Bond) of tissue slides, whole-slide imaging at 40x-resolution, and multiplexing of tissue sections from multiple patients within individual capture areas for spatial transcriptomics profiling. Using a cohort of thirteen pT3 stage colorectal cancer (CRC) patients, we compared the efficacy of deep learning models trained on slide prepared using an enhanced workflow as compared to the traditional workflow which leverages manual tissue staining and standard imaging of tissue slides. Leveraging Inceptionv3 neural networks, we aimed to predict gene expression patterns across matched serial tissue sections, each stemming from a distinct workflow but aligned based on persistent histological structures. Findings indicate that the enhanced workflow considerably outperformed the traditional spatial transcriptomics workflow. Gene expression profiles predicted from enhanced tissue slides also yielded expression patterns more topologically consistent with the ground truth. This led to enhanced statistical precision in pinpointing biomarkers associated with distinct spatial structures. These insights can potentially elevate diagnostic and prognostic biomarker detection by broadening the range of spatial molecular markers linked to metastasis and recurrence. Future endeavors will further explore these findings to enrich our comprehension of various diseases and uncover molecular pathways with greater nuance. Combining deep learning with spatial transcriptomics provides a compelling avenue to enrich our understanding of tumor biology and improve clinical outcomes. For results of the highest fidelity, however, effective specimen processing is crucial, and fostering collaboration between histotechnicians, pathologists, and genomics specialists is essential to herald this new era in spatial transcriptomics-driven cancer research.

## Introduction

For centuries, histological examination of tissue has been fundamental in disease prognostication [1]. Although such examination remains a cornerstone in pathology, the advent of genomic technologies has broadened our understanding of tumorigenesis, highlighting the value of examining expression patterns to gain comprehensive insights into tumor behavior and therapeutic response [2–4]. Typically, histopathological analysis is supplemented by immunohistochemical staining [5]. These evaluations provide spatial insights into molecular signatures that underscore cellular heterogeneity within a tissue sample. However, most immunohistochemical and fluorescence assays are limited in the number of markers they can analyze simultaneously. This limitation has been addressed with the emergence of spatial transcriptomics technologies, such as the Visium platform from 10x Genomics, which offers high multiplexing capability at remarkable spatial resolution, transforming our capacity to study expression patterns within intricate, nuanced tissue architectures [6,7]. When traditional histology is paired with spatial transcriptomics, it facilitates the correlation and combination of histopathological features with specific gene expression patterns. This integrative approach not only deepens our grasp of tumor heterogeneity and microenvironmental interactions but also holds the promise of uncovering novel spatial correlations that can refine prognostic evaluations.

As current spatial transcriptomics assays are costly, elucidating the connection between tissue histomorphology and underlying molecular signatures can help provide multiplexing capabilities at scale, which are often limited by cost and reproducibility [6]. Current methods for inference of spatial transcriptomic patterns are inspired by virtual staining techniques [8–10], which use computational methods to predict molecular traits from routinely collected histological images. This obviates the need for further tissue staining/assaying. Preliminary studies have supported the potential of these techniques to expand highly multiplexed spatial molecular evaluations to more extensive cohorts in a cost-effective manner. Yet, two primary obstacles impede consistent and high-quality spatial molecular inference from large, external sample groups using conventional stains. First, the quality of whole slide images must be elevated. Leveraging cutting-edge imaging technologies, such as the Aperio GT450s, coupled with the uniform application of staining reagents via automated slide staining machines, can pave the way for consistent results [11]. Given the necessity for accurately co-registered imaging and omics data, it’s posited that refined staining and advanced imaging might enable more detailed evaluations. Up to this point, most experiments aiming to derive expression from tissue have depended on manual staining techniques and lower-resolution imaging. Given these limitations, it’s unsurprising that the field has not yet realized the full potential or expanded the extensive range of biomarkers possible. Second, expanding the sample size of cohorts to train these virtual staining algorithms without escalating costs is imperative to capture a wider spectrum of patient and tumor attributes, ensuring a more thorough and reliable analysis with application to larger cohorts. Third, it is important to be able to allow for a much richer picture of the underlying relationship between immune cells and tumor tissue.

The recent introduction of the CytAssist device potentially addresses these concerns [12,13]. In contrast to the traditional Visium FFPE assay, which mandates manual staining and specific Visium slide imaging conditions (loosely adhered coverslips and short imaging window), the CytAssist allows for the stable coverslipping of slides and extended time frame between staining, imaging, and analyte retrieval. In addition, the CytAssist workflow relies on tissue sections placed onto standard histology slides rather than costly Visium barcoded slides, simplifying tissue placement and allowing the selection of specific ROIs for analysis. Together, this innovative design allows multiple tissue sections to be amalgamated onto a single slide before Visium profiling and facilitates the utilization of automated staining technologies and cutting-edge imaging using clinical-grade pathology infrastructure. These improvements not only augment whole slide image quality for intricate, deep learning analyses but might also considerably diminish associated costs.

In our research, we demonstrate some of the advantages brought by the CytAssist device in spatial evaluations. By multiplexing two tissue samples onto one slide, cost reductions become tangible. Incorporating automated staining procedures and superior imaging modalities enhances whole slide image quality, paving the way for more precise, in-depth analyses. Here, we focus on the effective integration of multiple tissue samples via the CytAssist tool, showcasing its potential to elevate spatial expression inferences from standard H&E stains when juxtaposed against manual staining and inferior imaging of analogous capture regions from sequential sections.

Our comparison centers around a cohort of colorectal cancer patients. Colon cancer, increasingly affecting younger age groups, is a major global health issue due to its high prevalence and mortality. Crucial for determining prognosis and guiding treatment, colon cancer staging primarily relies on the TNM classification, which evaluates tumor invasion, lymph node involvement, and metastasis [14–17]. Notably, metastasis critically affects patient outcomes, marking increased tumor aggressiveness. While the TNM system is pivotal, it might not encompass the entire complexity of tumor biology, prompting research into additional molecular markers to enhance prognostic accuracy and predict recurrence risks [18]. Discovering new biomarkers from the spatial distribution of specific genes within the tumor and its immune microenvironment can provide insights into antitumoral reactions, potentially enhancing colon cancer staging [19–21].

## Methods

### Data Collection

#### Specimen Overview

Our dataset comprises specimens processed through two distinct workflows: the traditional workflow (4 patients, 4 capture areas) and the new enhanced workflow (8 patients, 4 capture areas). In addition, two paired serial sections were profiled for comparative analysis, one emulating the traditional workflow and the other the new workflow, both leveraging the CytAssist technology.

#### Patient and Capture Area Selection

This dataset represented thirteen patients diagnosed with pathologic T Stage-III (pT3) colorectal cancer. These patients were selected through a retrospective review of pathology reports from 2016 to 2019. Four patients were featured in a previous study where we restricted these patient characteristics based on microsatellite stable tumors and tumor site (right/transverse colon) [22]. For the remaining cohort of nine patients, to ensure a balanced representation of patient characteristics, the patients were matched based on various criteria, including age, sex, tumor grade, tissue size, and mismatch repair/microsatellite instability (MMR/MSI/MSS) status [23]. MSI status was determined by assessing the loss of expression of MLH1 and PMS2 through immunohistochemistry. Tissue blocks were sectioned into 5-10-micron thick layers, and specific regions of interest such as epithelium, tumor-invasive front, intratumoral areas, and lymphatics. Capture areas were annotated by a pathologist from serial whole slide images (WSI). Representative regions were carefully dissected from the tissue, placed into capture areas, and subjected to H&E staining, imaging, and Visium profiling in the Pathology Shared Resource at the Dartmouth Cancer Center and Single Cell Genomics Core in the Center for Quantitative Biology.

#### Traditional Workflow

The first four capture areas (Capture Areas 1-4; **Table 1, Figures 1A,2**) were profiled using the traditional 10x Visium workflow– after macrodissection, placement onto the Visium barcoded slide, and manual H&E staining, the Visium workflow: 1) images the tissue at 10-20x resolution using standard image scanning (EVOS m7000 scanner, Thermo Fisher), 2) the tissue is permeabilized for hybridization of whole transcriptome mRNA probes, followed by, 3) probe ligation and release for capture on the Visium slide through poly(A) tail binding; 4) next, captured probes are extended and amplified to incorporate spatial barcodes and 5) the probes and spatial barcodes are sequenced on an Illumina NovaSeq instrument [24] targeting 50,000 reads/spot. The 10x Genomics SpaceRanger software is used to convert raw sequencing data into spatially-resolved gene expression matrices. This comprehensive process enables whole transcriptome (mRNA) profiling of up to 5,000 55uM spots with a 100uM center-to-center distance within a 6.5 mm^2^ capture area or 14,000 spots within an 11 mm^2^ capture area. After post-filtering uninformative reads, we obtained approximately 17,943 genes at nearly 5,000 locations for each slide (total Visium spots: 4950, 4922, 4887, and 4169 per slide).

**Table 1:** Description of Patient Cohort. Tissue sections from 13 patients (14 sections) are divided amongst 10 capture areas, with up to two tissue sections per capture area joined together (left/right side). **\***Indicates held-out capture areas 5 and 6 from serial sections from the same patient and location for testing. All other tissue sections come from different patients.

| Workflow | Capture Area | CytAssist | Dimension (mm^2) | Section Placed on Left Side / Center of Capture Area |  |  |  |  | Section Placed on Right Side of Capture Area |  |  |  |  |
| --- | --- | --- | --- | --- | --- | --- | --- | --- | --- | --- | --- | --- | --- |
|  |  |  |  | Age | Sex | Tumor Site | MSI Status | METS | Age | Sex | Tumor Site | MSI Status | METS |
| Traditional: Manual Stain + Low Resolution Imaging | 1 | no | 6.5x6.5 | 40-45 | F | Transverse Colon | MSS | yes | – | – | – | – | – |
|  | 2 | no | 6.5x6.5 | 60-65 | M | Right Colon | MSS | no | – | – | – | – | – |
|  | 3 | no | 6.5x6.5 | 80-85 | M | Right Colon | MSS | yes | – | – | – | – | – |
|  | 4 | no | 6.5x6.5 | 45-50 | F | Right Colon | MSS | no | – | – | – | – | – |
|  | 5* | yes | 6.5x6.5 | 80-85 | M | Left Colon | MSI | yes | – | – | – | – | – |
| Enhanced: Automated Staining + High Resolution Imaging | 6* | yes | 6.5x6.5 | 80-85 | M | Left Colon | MSI | yes | – | – | – | – | – |
|  | 7 | yes | 11x11 | 90-95 | F | Hepatic Flexure | MSI | yes | 80-85 | M | Left Colon | MSI | no |
|  | 8 | yes | 11x11 | 85-90 | F | Splenic Flexure | MSS | yes | 70-75 | F | Hepatic Flexure | MSS | no |
|  | 9 | yes | 11x11 | 80-85 | F | Cecum | MSI | yes | 70-75 | M | Cecum | MSI | no |
|  | 10 | yes | 11x11 | 55-60 | F | Left Colon | MSS | yes | 65-70 | M | Sigmoid | MSS | no |

**Figure 1:**
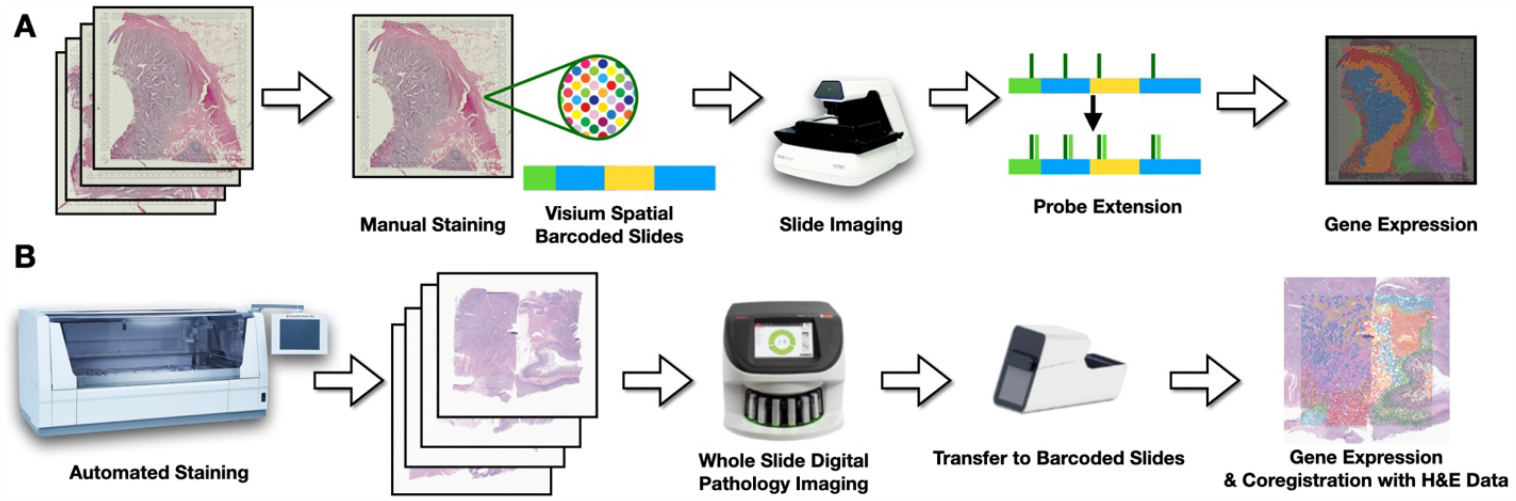
Comparative Overview of the Two Workflows. **(A)** Traditional workflow: After placing tissue on Visium barcoded slide, sections are manually stained with H&E and imaged using the EVOS m7000. **(B)** Enhanced workflow: Automated application of chemical reagents with 40x resolution imaging via Aperio GT450, followed by transfer to Visium device facilitated by 10X CytAssist.

**Figure 2.**
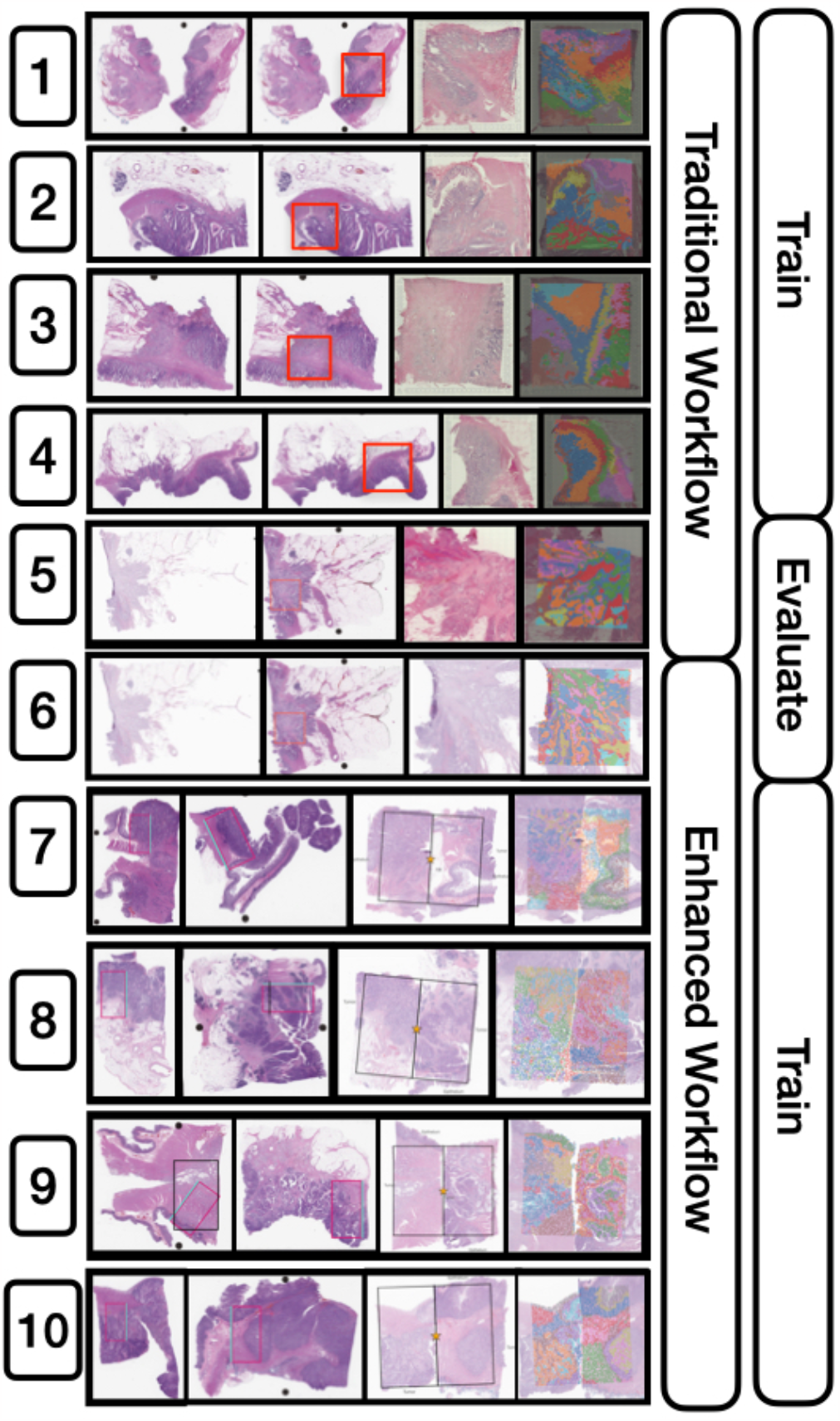
Detailed Configuration of the 10 Capture Areas and Their Preprocessing Workflow. The figure presents the progression of all 10 capture areas, from their inception to visualization. The sequence is as follows: 1) H&E-stained slide, 2) Selected capture area dedicated to one or two patients, 3) Visualization of capture area, 4) Integration with spatial transcriptomics clusters. Training involved the use of either traditional, enhanced, or a combination of both slide types. Notably, capture areas 5 and 6 were set aside as held-out serial sections for subsequent analyses.

#### Enhanced Workflow

We trialed an improved specimen processing workflow designed specifically for profiling specimens with the Visium CytAssist assay. This protocol integrates the CytAssist technology with improved specimen processing in Pathology to ensure consistent staining and optimal image quality by leveraging the capabilities of pathology department automated slide stainer via the Sakura Tissue-Tek Prisma Stainer (Sakura Finetek USA, Inc. 1750 West 214th Street, Torrance, CA 90501) [25] and the Aperio GT450s imaging system. The imaging was conducted at a high-resolution of 40x (equivalent to 0.25 micron per pixel) before proceeding with Visium profiling. Four tissue slides were collected representing eight patients, resulting from macrodissection of tissue sections from FFPE blocks. Patient selection criteria were well matched to the set used for the traditional workflow save for microsatellite instability status and age, which featured additional variation in this expanded set of patients. These sections were precisely marked by pathologists in serial WSIs to target specific tissue architectures. To maximize resource efficiency, tissue segments from two patients were merged onto a single slide, creating an 11mm by 11mm capture region. This strategy ensured each capture area contained an equal representation of metastasis and MSI status from anatomically similar sites. Using our improved workflow, we first 1) placed FFPE tissue sections onto standard histology slides, followed by coverslipping in a glycerol + xylene mounting medium, 2) performed deparaffinization, rehydration and H&E staining on a Leica Bond instrument, 3) collected whole slide images at 40x resolution on Aperio GT450 scanners, and 4) decoverslipped in xylene for 1-3 days (until coverslips were detached). The remaining steps of destaining, probe hybridization, probe ligation, eosin staining, transfer to the Visium slide using CytAssist, and library preparation were performed according to the manufacturer’s protocol (CG000485). Libraries were sequenced on an Illumina NovaSeq targeting 50,000 reads/spot. This detailed method permits unbiased gridded profiling of spots within slides area. The subsequent imaging of the same tissue slide (after staining with eosin) facilitated precise co-registration of the 40X high-resolution pathology slide with the Visium spatial transcriptomics. After the manual selection of fiducials, the Spaceranger software was employed to align CytAssist sections with their corresponding 40X H&E stains, which ensures accurate co-registration, and conduct quality control and convert the Visium Spatial Transcriptomics (ST) data into an easily interpretable format (**Table 1, Figures 1B,2**). It should be noted that this enhanced workflow does not yet apply to fresh frozen sections.

#### Comparison Slides

To mitigate the influence of inherent tissue variability and rigorously assess the CytAssist technology, we designated two slides as comparison points to evaluate our machine learning models. These were based on paired serial sections spatially matched to represent identical capture areas. Capture Areas 5 and 6 were earmarked for our comparative analysis of the CytAssist technology (**Table 1, Figure 2**). Each slide from these areas underwent distinct preparation methods to mirror both the traditional and our improved workflow. From these slides, the set of nearly 18,000 genes was reduced to the 1,000 most spatially variable genes using the SpatialDE package [26].

### Machine Learning Modeling and Evaluation

For the enhanced workflow, co-registered 40X WSI corresponding to each Visium slide were cropped into 512-by-512 pixel sub-images centered around each Visium spot within the capture areas, selected based on a previous study that had conducted a sensitivity analysis over various patch sizes [22]. For the traditional workflow, each Visium spot encompasses a circular capture zone with a 130-pixel diameter at a 20x magnification. Several deep learning models– with an Inceptionv3 convolutional neural network architecture as described in previous work– were trained to predict Visium ST at each spot for both binary (i.e., low/high expression, dichotomized by median expression) and continuous (e.g., log-transformed of pseudo counts with an offset of 1 read) prediction tasks for 1,000 spatially-variable genes. Models were trained using the mean squared error on the log-transformed counts for continuous data. As aforementioned, performance for dichotomous tasks was calculated through dichotomization via median expression [27,28]. Model parameters were selected based on optimal performance statistics across an internal validation set across the training epochs. Hyperparameters were set based on a coarse hyperparameter search for each method.

### Experimental Comparisons

We used a comprehensive comparative analysis to discern the benefits of the enhanced workflow against the traditional workflow. The experimental comparisons probed the workflows under various training and validation regimes, thereby providing insights into their relative strengths and potential synergies (**Table 2**). We evaluated our models using paired serial sections from both workflows, ensuring minimal tissue variability to highlight the impact of staining and imaging methods. The performance comparisons were based on the training sets described in **Table 2**.

**Table 2:** Comparative Analysis of Model Training Approaches Across Distinct Workflows. This table delineates the methodologies used for generating training data from both the traditional and enhanced workflows. Emphasis is placed on evaluating the implications of different staining and imaging methods. By employing reserved comparison samples, models trained on each dataset undergo assessment using paired serial sections from both workflows, aiming to reduce tissue-related variability.

| Training Data | Purpose | Predictive Analysis |
| --- | --- | --- |
| Traditional Workflow Slides | To establish a foundational performance baseline. | This comparison gauged the ability of models trained on the traditional method to predict spot-level expression across both techniques using the paired serial sections (encompassing both traditional and improved workflows). |
| Enhanced Workflow Slides | To spotlight the enhanced capability and superiority of the improved workflow. | Evaluating predictions on the paired serial sections from both workflows showcases how models, when trained sections assayed through the improved workflow, interpret results from both techniques. Ideally, its performance should meet or surpass the traditional workflow's metrics on the shared paired slides. |
| Slides from Both Workflows | To harness the collective merits of both workflows, forging a holistic understanding of tissue histology across various specimen processing and imaging methods. | Training on tissue acquired from both protocols promises to impart a broader representation of the data to the models. Evaluations on the comparison slides reflect the model's adaptability, informed by both workflows. |

### Performance Evaluation

#### Evaluative assessments on comparison slides

Spot level expression was compared on the held-out comparison slides, retaining their native imaging resolutions. Direct evaluation focused on recapitulating the expression at each individual Visium spot across the entirety of the held-out slides. This granular assessment ensured an in-depth understanding of how well the trained models can predict expression profiles at localized regions throughout the slides. Confidence in model performance is reported through 95% confidence intervals derived from 1000-sample non-parametric bootstrapping of Visium spot observations.

#### Performance Metrics

Performance metrics include: **Quantitative Metrics**, Area Under the Receiver Operating Characteristic Curve (AUC) for dichotomous tasks and Spearman correlation for continuous tasks, macro-averaged across all genes. **Qualitative Evaluation**, beyond quantitative scores, we examined the capability of each approach to mirror true expression patterns. This involved comparing the clustering of true expression patterns on those predicted from the tissue histology, utilizing the AlignedUMAP dimensionality reduction technique to generate visually comparable low-dimensional embeddings [29,30]. A more effective method should ideally maintain the uniqueness and structure of the original clusters.

#### Differential Expression

As a final comparison between the traditional and enhanced workflow, we aimed to gauge the predictive accuracy of each method in relation to tissue architecture. Specifically, each Visium spot was annotated according to its location—either within the tumor, at its periphery, or distal to the tumor. We hypothesized that the enhanced slides would yield gene expression profiles more reflective of these distinct tissue regions, approximating the precision of the ground truth expression more closely than the traditionally processed slides would. To test this, we employed the Mann-Whitney U-test to analyze differential gene expression (treating expression as a continuous count-based measure) between the tumor-interface zones and those regions either within or away from the tumor [31]. This analysis focused on the top-200 genes as ranked by the Spearman correlation statistics between the true and predicted expression. We then compared the U-statistics obtained from the actual expression data to those generated from predicted expression, summarizing the results as the median percentage change in U-statistics across the examined genes, with 95% confidence intervals reported using 1000-sample non-parametric bootstrapping.

## Results

### Enhanced Staining and Imaging Workflow Results in Substantial Boost in Predictive Performance

When assessing the predicted expression against the true expression for the top 1000 spatially variable genes in the held-out slides, the models demonstrated remarkable accuracy, reported using both AUC and Spearman statistics (**Table 3, Figure 3, Supplementary Figure 1**).

**Table 3:**
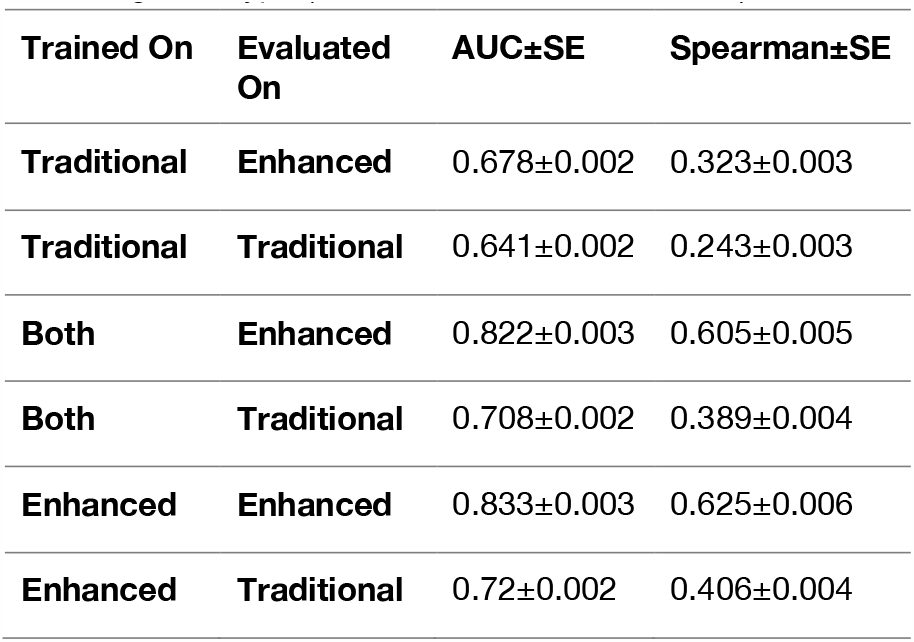
Performance Metrics for Held-Out Capture Areas Across Top 1000 Genes. This table presents the median AUC and Spearman correlation coefficients and their respective 95% confidence intervals derived from a 1000-sample non-parametric bootstrap. Metrics are shown for each combination of training slide type (traditional, enhanced, or both) and evaluation slide type (traditional or enhanced).

| Trained On | Evaluated On | AUC±SE | Spearman±SE |
| --- | --- | --- | --- |
| Traditional | Enhanced | 0.678±0.002 | 0.323±0.003 |
| Traditional | Traditional | 0.641±0.002 | 0.243±0.003 |
| Both | Enhanced | 0.822±0.003 | 0.605±0.005 |
| Both | Traditional | 0.708±0.002 | 0.389±0.004 |
| Enhanced | Enhanced | 0.833±0.003 | 0.625±0.006 |
| Enhanced | Traditional | 0.72±0.002 | 0.406±0.004 |

**Figure 3:**
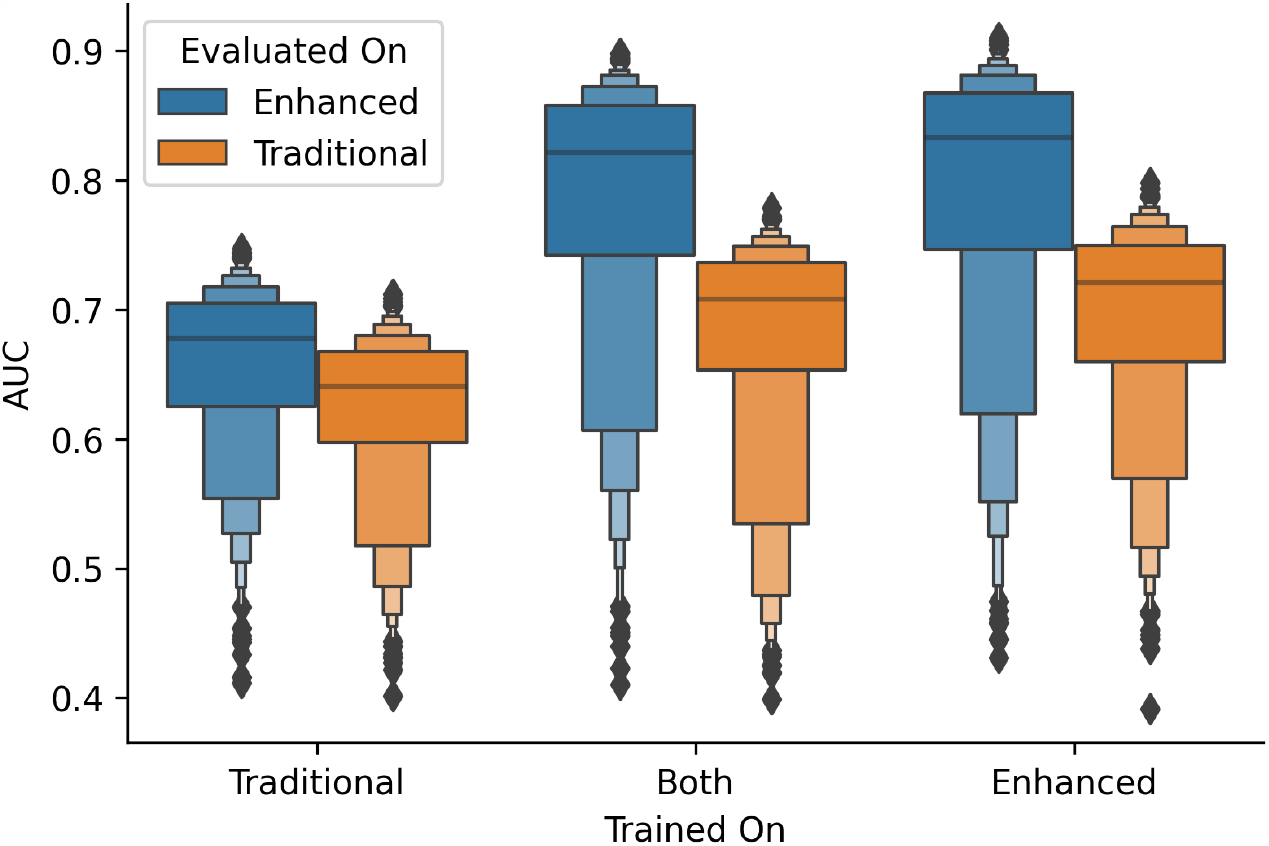
Boxenplot of AUC Performance Across Top 1000 Genes. This plot showcases the comparative performance of held-out capture areas based on training slide type (traditional, enhanced, or both) and evaluation slide type (traditional or enhanced), using the area under the receiver operating characteristic curve as the performance metric.

Overall, models trained using the traditional dataset predicted expression on both traditional and enhanced slides with approximately 0.66 AUC and a 0.28 correlation. These models were notably the underperformers. In contrast, exclusively leveraging the enhanced workflow led to a major increase in predictive performance. Specifically, while training and testing on traditional slides yielded an AUC of 0.641 and a 0.243 correlation, the same process on enhanced 40x-resolution WSI (enhanced workflow) catapulted the results to an AUC of 0.833 and a 0.625 correlation—this translates to a surge of nearly 45% in AUC and a staggering 157% in Spearman correlation. Our exploration into whether a hybrid training approach, incorporating both traditional and enhanced slides, would augment performance turned out to be inconclusive, as it did not notably elevate predictive power for either slide type and instead lead to modest reductions in performance from training solely on WSI acquired through the improved workflow. As depicted in **Figure 4**, visually, the expression patterns across a slide appear more clearly distinct and align more closely with the ground truth when training is conducted using enhanced slides, irrespective of whether the comparison slide utilized enhanced or traditional techniques.

**Figure 4:**
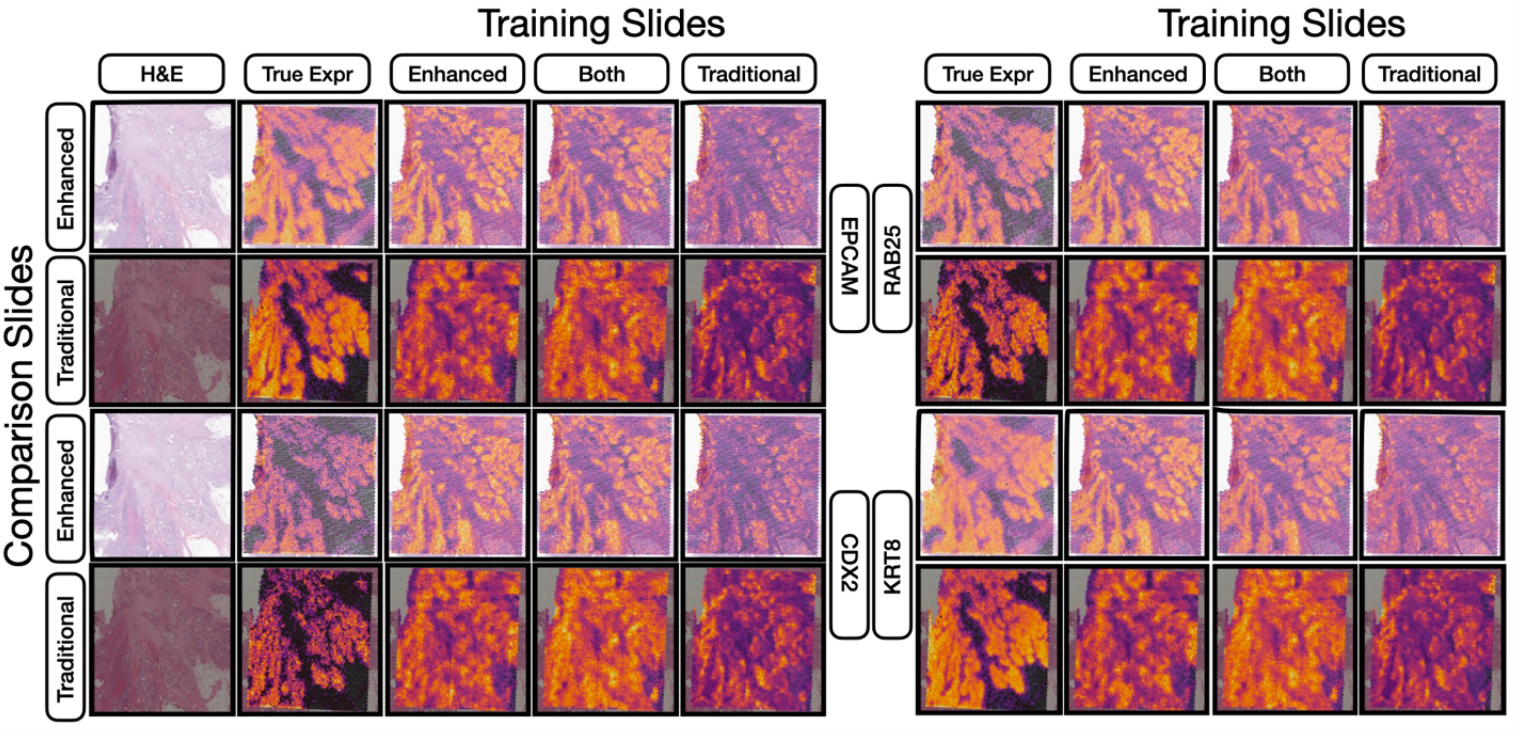
Heatmap Visualization of Gene Expression Predictions. This figure juxtaposes the ground truth gene expression heatmaps against the predictions made by neural networks trained on either enhanced slides, both preparation approaches or traditional slides. Each prediction is showcased for both comparison slide types (traditional and enhanced). Specific markers from the held-out slides (both enhanced and traditional) are highlighted to emphasize the nuanced differences across training techniques and their evaluation.

### Topological Consistency

Similar to previous works [22,27], we sought to understand whether slides processed using the enhanced workflow yielded predicted expression patterns that were more topologically consistent with the ground truth expression, i.e., similarly clustered. Overlaying the clustered ground truth Visium spot expression on the predicted embeddings would provide a subjective measure of which methods provide better clustering. This topological agreement was compared for predicted expression patterns from models utilizing various combinations of enhanced and traditional slides for training. For evaluations conducted on the enhanced WSI, models trained using both enhanced and traditional slides together, as well as those trained solely on enhanced slides, produced the most pronounced clustering (**Figure 5**). The relative placements of these clusters closely reflected the ground truth. Conversely, the resulting clusters were less defined when models were assessed on the traditional slide. Notably, models trained on traditional slides seemed to conflate multiple clusters.

**Figure 5:**
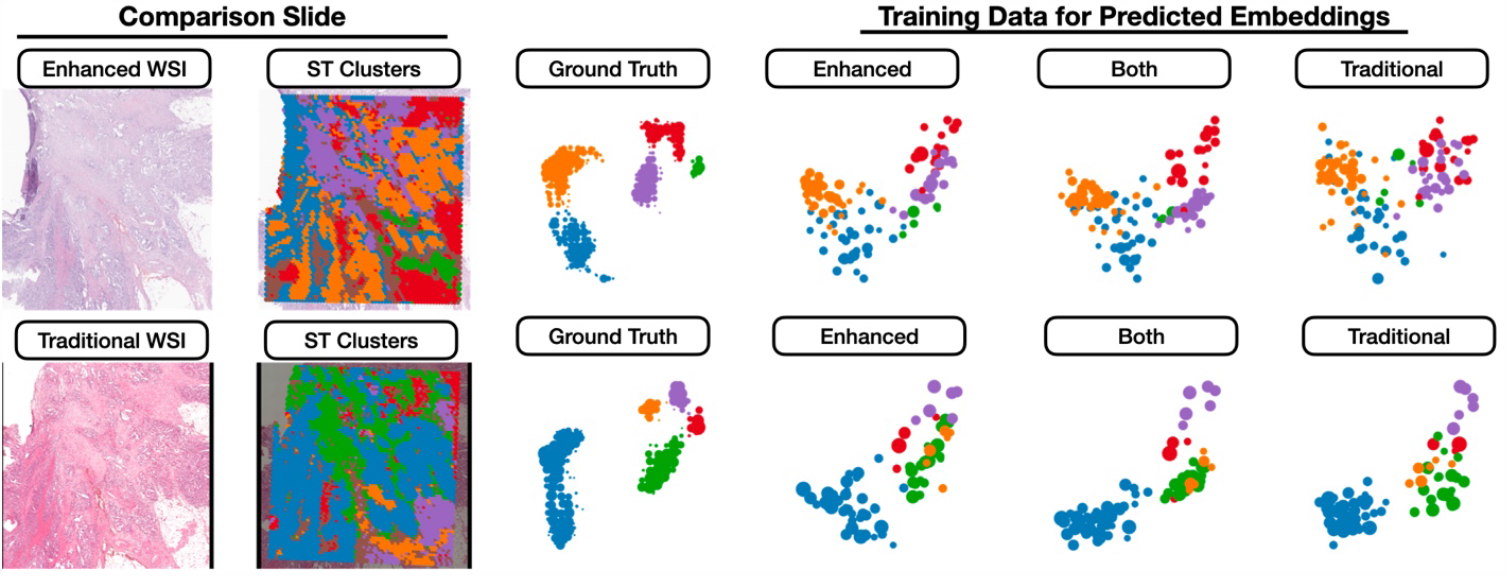
UMAP embeddings of true and predicted gene expression for enhanced and traditional evaluation slides. The Mapper algorithm was employed to flexibly group together embeddings, with groups containing overlapping Visium spots. Node sizes in the Mapper plots correspond to the number of Visium spots, and colors indicate the dominant cluster membership. The clusters were assigned to each visium spot using AlignedUMAP embeddings followed by HDBSCAN clustering on the ground truth count-based expression data.

### Differential Expression and Prediction of Tissue Architecture

We evaluated each method’s capability to predict expression patterns characteristic of the tumor invasive margin in contrast to regions inside and distant from the tumor. Given the potential variability in predicted expression scales, we employed the Mann-Whitney U test to contrast expression across these tissue structures, documenting the percentage shift in U-statistics between actual and predicted expression. These findings are consistent with our predictive performance observations, suggesting that the ability to accurately predict expression is synonymous with more refined delineations of tissue architecture. This predictive performance and precision in the subsequent differential expression analyses are notably enhanced by the CytAssist leveraging the enhanced workflow (**Table 4**). For example, models either trained on both enhanced and traditional slides or exclusively on enhanced slides were most successful in recapitulating the U-statistics derived from actual expression data when assessed on enhanced slides. However, there was a marked drop in accuracy when these metrics were applied to the reserved traditional slide.

**Table 4:** Comparative Performance on Tumor Interface Markers Localization: This table showcases the percentage variations in U-statistics between true and predicted gene expression (continuous count data) for the top-200 genes, emphasizing the method’s precision in identifying expression differences at the tumor invasive margin relative to both its internal and surrounding areas. 95% confidence intervals were derived from a 1000-sample non-parametric bootstrap to measure the robustness of these findings. A diminished percentage difference signifies a heightened capability to pinpoint molecular markers at the tumor interface in a manner similar to the actual expression.

| Trained On | Evaluated On | U-Statistic Percent Change (%) | 2.5% CI | 97.5% CI |
| --- | --- | --- | --- | --- |
| Traditional | Enhanced | 18.73 | 18.42 | 18.93 |
| Traditional | Traditional | 14.19 | 13.79 | 14.43 |
| Both | Enhanced | 5.23 | 4.80 | 5.61 |
| Both | Traditional | 17.56 | 17.33 | 17.84 |
| Enhanced | Enhanced | 5.40 | 5.22 | 5.77 |

| Enhanced | Traditional | 17.11 | 16.90 | 17.47 |
| --- | --- | --- | --- | --- |

## Discussion

Our study aimed to evaluate the influence of specimen processing in the realm of spatial transcriptomics and spatial transcriptomics inference. Here, we compared traditional and enhanced workflows for the task akin to virtual staining—a technique that can infer spatial expression patterns directly from whole slide image histomorphology [8,32]. This method offers the potential to democratize spatial transcriptomics insights to more extensive cohorts for genes exhibiting high predictability, subsequently broadening the spectrum of markers under consideration. Enhancements in specimen preparation, specifically in three key areas including: 1) tissue multiplexing to reduce costs based on their positioning in the mounting medium, 2) optimizing staining procedures, and 3) refining imaging processes, could facilitate more accurate image-based RNA inference and other integrative analysis, thereby boosting statistical precision. The incorporation of CytAssist was pivotal, offering insights into how upstream enhancements in specimen processing can yield vastly improved *in silico* outcomes.

### Principal findings in the context of improved tissue staining

This study underscores the discernible performance variations between slides processed through enhanced and traditional workflows, reflecting differences in tissue staining and imaging in the context of deep learning applications for spatial transcriptomics [33]. Tissue staining, a technique to enhance the contrast between various tissue components, is of paramount importance. Dyes such as hematoxylin and eosin, with their distinct optical absorption properties, offer a range of color variations. When oxidized, hematoxylin interacts with various metals, forming complexes that produce unique colors, enhancing the dye’s staining capabilities [33–41]. Even minor deviations in staining procedures and timing can result in fluctuating staining intensity. Human variations in the timing of staining and the use of reagents nearing their expiration or when overused/over-oxidized/deteriorated can further compromise quality. Factors such as contaminants also introduce inconsistencies, impacting the uniformity of tissue staining. Automated staining solutions offer a promising alternative to manual methods, eliminating human-induced sources of variation. By using software to control the application of H&E stains according to a set protocol, both the quality and consistency of specimens can be enhanced. Removing these variations allows algorithms to shift their focus from capturing variability to representing the underlying structures with greater fidelity [35]. Past research validates that digital image analysis is frequently compromised by inconsistent staining. However, automating this process has been shown to not only improves staining consistency but also bolsters the contrast in tissue structures, thereby increasing diagnostic reliability [33–41]. For algorithms to effectively discern gene-related histologies, consistently capturing intricate details, better represented by minimizing these sources of variability, is vital.

Thus, it is unsurprising that tissues stained through automated processes exhibited superior performance. The models trained on tissue sections processed using the enhanced workflow demonstrated remarkably stronger predictive accuracy, as evidenced by higher AUC and Spearman correlation values. Evaluation of these models on enhanced slides also presented a performance advantage, even when only training on traditionally processed slides. Moreover, we showcased that heightened predictive accuracy can lead to a bolstered statistical prowess in evaluating tissue architecture with enhanced slides relative to their traditional counterparts. Analysis of the aligned UMAP embeddings demonstrated that enhanced slides tended to yield expression patterns that were more topologically consistent with the ground truth, indicating their potential to cluster data more effectively.

### Interpretation of Findings and the Need for Broader Validation

Our research affirms that by prioritizing specimen preparation and imaging, especially with the aid of CytAssist, one can amplify the statistical acuity of subsequent analyses and more authentically capture the intricate relationships among Visium spots from histological observations [42,43]. This heightened precision, made possible by the enhanced staining and imaging workflow, has the potential to illuminate the molecular intricacies and spatial configurations of unique tissue structures. Such insights pave the way for a more profound comprehension of CRC metastasis, especially when these state-of-the-art techniques are applied to broader cohorts.

### Challenges and Future Directions

This study focused on comparing enhanced and traditional workflows within a specific set of capture areas. The derived insights offer a foundational framework for both validating and scaling these techniques to expansive cohorts. However, there are a few considerations which warrant further attention. Firstly, the comparison between traditional and enhanced workflows was limited to a specific set of capture areas, necessitating further exploration to broaden the application, scope, and impact of our findings. Generalizing our findings to other tissue types, molecular pathways, and experimental setups should be further explored. For instance, the enhanced workflow does not yet apply to fresh frozen sections which will be the subject of future work. To affirm the universality and adaptability of the models, varied staining methodologies, slide preparations, and tissue specimens should be considered, requiring additional forms of validation (e.g., immunostaining, alternative spatial transcriptomic assays) [44–46]. Such disparities can introduce unpredicted variability, with potential ramifications on model efficiency. Although there are algorithmic solutions for standardizing staining agents, a holistic approach may require a collaborative multicenter framework, strategies to alleviate batch inconsistencies, and close coordination amongst key stakeholders within various shared resource infrastructures across each institution [47].

Enhancing the scope of validation for deep learning paradigms as well as identifying areas for improvement outside of algorithmic development (e.g., specimen processing), can catalyze the more widespread integration of these nascent spatial transcriptomics technologies. The relevance of these findings hinges on external validation through independent cohorts. Moreover, the implications of our study should be expanded to encompass other diseases that warrant spatial molecular assessments [48–50].

## Conclusion

The validation of spatial transcriptomics information inferred from whole slide images provides a unique opportunity to assess colorectal cancer (CRC) metastasis with greater statistical precision. Conventional statistical analyses frequently rely on large volumes of specimens to yield significant results, which can prove costly for spatial transcriptomic assays. Yet, the spatial molecular data inferred through our advanced techniques, enhanced through sophisticated specimen processing, potentially diminish the necessity for such expansive and expensive datasets. Accurate extrapolations of gene expression landscapes within tissue samples can enable a more refined and purpose-driven exploration of CRC metastasis, further emphasizing our imperative to further validate these approaches in larger cohorts.

## Data Availability

Access to manuscript data is limited due to patient privacy concerns. All data produced in the present study are available upon reasonable request. Requests should be directed to senior author Dr. Joshua Levy.

## Acknowledgements

Human Research Protection Program IRB of Dartmouth Health gave ethical approval for this work. This study was carried out in the Genomics and Molecular Biology Shared Resource (GMBSR) at Dartmouth which is supported by NCI Cancer Center Support Grant 5P30CA023108 and NIH S10 (1S10OD030242) awards. Additionally, single cell genomics projects should include the following text (or similar): “Single cell studies were conducted through the Dartmouth Center for Quantitative Biology in collaboration with the GMBSR with support from NIGMS (P20GM130454) and NIH S10 (S10OD025235) awards. JL is funded under NIH subawards P20GM130454 and P20GM104416.

## Key points

This study showcases an enhanced workflow for deep learning-based spatial transcriptomics inference, using the flexibility of the Visium CytAssist assay to leverage improved tissue processing and imaging. Leveraging Inceptionv3 neural networks on slides from thirteen pT3 stage colorectal cancer patients, the enhanced method significantly outperformed traditional workflows in predicting gene expression patterns. The enhanced approach yielded gene expression profiles more topologically consistent with actual data, improving the statistical precision in identifying biomarkers linked to metastasis and recurrence. Effective specimen processing is paramount for high-fidelity results in spatial transcriptomics-driven cancer research. Working together, histotechnicians, pathologists, and genomics specialists play a critical role in advancing tissue preparation and imaging techniques, deepening our insight into tumor biology for further prognostic biomarker development.

## Author Biographical Information

1. **Michael Y. Fatemi*:** Undergraduate at the University of Virginia, specializing in computer science. Authored multiple publications on spatial transcriptomics inference from whole slide images.
2. **Yunrui Lu*:** Research assistant at Dartmouth Health with a data science focus. Expertise lies in applying deep learning to various digital pathology tasks.
3. **Alos B. Diallo:** Doctoral student at Dartmouth College, delving into the study of tumor microenvironments and cellular interactions.
4. **Gokul Srinivasan:** Research assistant at Dartmouth Health with a data science focus. Has led/contributed to several works on spatial transcriptomics inference from whole slide images.
5. **Zarif L. Azher:** High school student at Thomas Jefferson School for Science and Technologies. Known for his work on self-supervised methods for multimodal prognostication models.
6. **Brock C. Christensen:** Professor at Dartmouth College, directing research towards epigenetic profiling of tissues.
7. **Lucas A. Salas:** Assistant Professor at Dartmouth College, pioneering research on DNA methylation for cell typing in tumor immune microenvironments.
8. **Gregory J. Tsongalis**: Director of the CGAT Laboratory for Clinical Genomics and Advanced Technologies at Dartmouth Health. Well known for his comparative research on novel molecular assays.
9. **Scott M. Palisoul:** Expert histotechnician at Dartmouth Health who played a significant role in establishing innovative specimen preparation and image scanning protocols.
10. **Laurent Perreard:** Lab manager at Dartmouth College facilitating usage of the 10x Genomics CytAssist assay.
11. **Fred W. Kolling IV:** Director of the Single Cell Genomics Core at the Center for Quantitative Biology. Recognized for trialing novel spatial and single-cell transcriptomic technologies.
12. **Louis J. Vaickus:** Director of Medical Informatics in the Department of Pathology at Dartmouth Health. Blends pathologist and AI expertise.
13. **Joshua J. Levy**:** Co-director of the EDIT AI group and director of digital pathology research at Cedars Sinai Medical Center. Specializes in studying spatial molecular heterogeneity of tumors and developing diagnostic technologies.

## Supplementary Materials

**Supplementary Figure 1:**
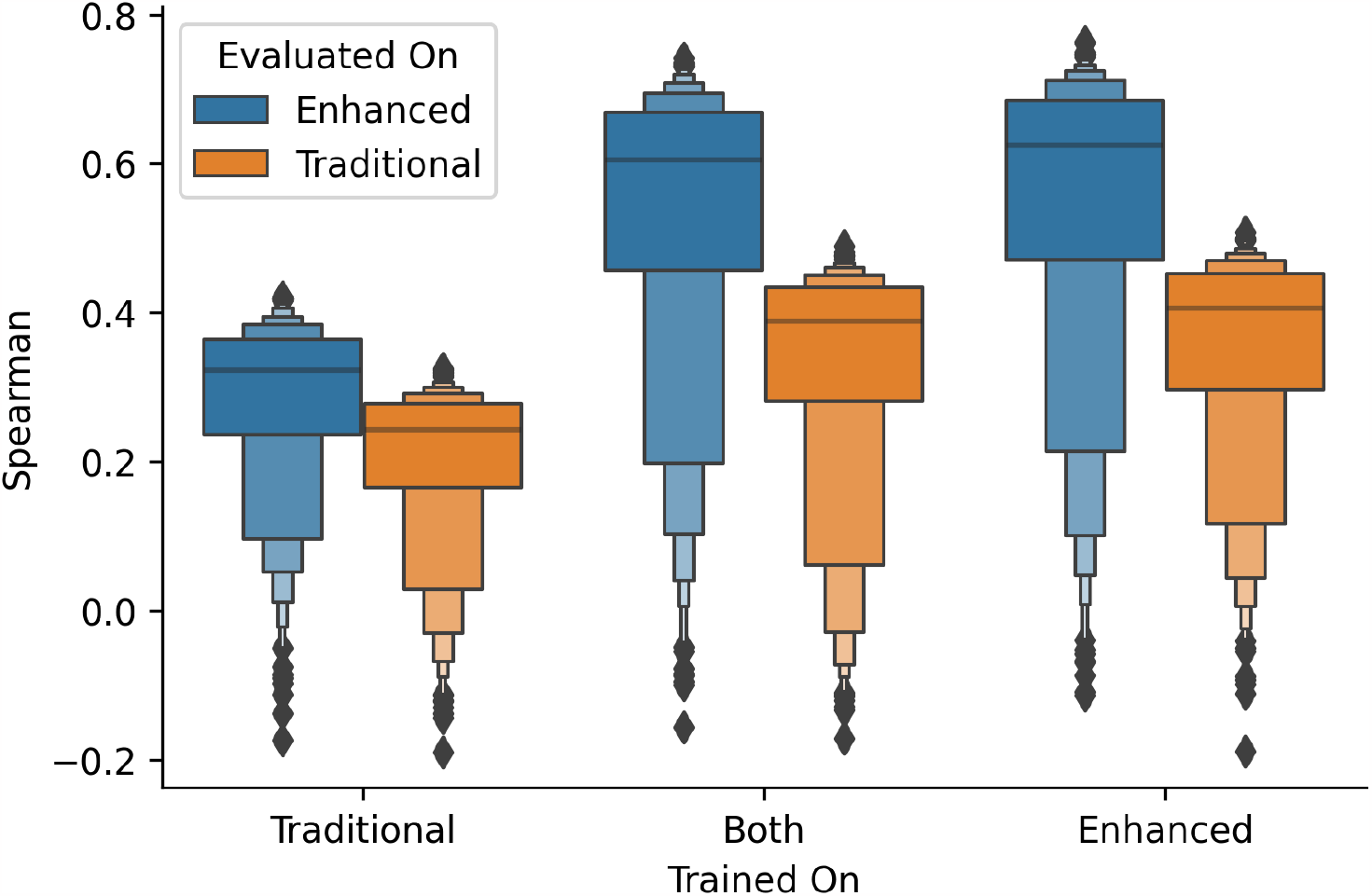
Boxenplot of Spearman Performance Across Top 1000 Genes. This plot showcases the comparative performance of held-out capture areas based on training slide type (traditional, enhanced, or both) and evaluation slide type (traditional or enhanced), using the Spearman correlation coefficient as the performance metric.

## Notes

### Competing Interest Statement

The authors have declared no competing interest.

### Author Declarations

Human Research Protection Program IRB of Dartmouth Health gave ethical approval for this work.

